# HYDROXYCHLOROQUINE FOR THE TREATMENT OF SEVERE RESPIRATORY INFECTION BY COVID-19: A RANDOMIZED CONTROLLED TRIAL

**DOI:** 10.1101/2021.02.01.21250371

**Authors:** Carmen Hernandez-Cardenas, Ireri Thirion-Romero, Norma E. Rivera-Martinez, Patricia Meza-Meneses, Arantxa Remigio-Luna, Rogelio Perez-Padilla, on behalf of the Research Group on hydroxychloroquine for COVID-19

## Abstract

The novel coronavirus pandemic (COVID–19) represents a major public health problem due to its rapid spread and its ability to generate severe pneumonia. Thus, it is essential to find a treatment that reduces mortality. Our objective was to estimate whether treatment with 400 mg/day of Hydroxychloroquine for 10 days reduces in-hospital mortality in subjects with severe respiratory disease due to COVID-19 compared with placebo.

**Material and methods:** A double-blind, randomized, placebo-controlled trial to evaluate the safety and efficacy of Hydroxychloroquine for the treatment of severe disease by COVID-19 through an intention-to-treat analysis. Eligible for the study were adults aged more than 18 years with COVID-19 confirmed by RT-PCR and lung injury requiring hospitalization with or without mechanical ventilation. Primary outcome was 30-day mortality. Secondary outcomes: days of mechanical ventilation, days of hospitalization and cumulative incidence of serious adverse events.

**Results:** A total of 214 patients with COVID-19 were recruited, randomized and analyzed. They were hypoxemic with a mean SpO_2_ of 65% ± 20, tachycardic (pulse rate 108±17 min-^1^) and tachypneic (32 ±10 min-^1^); 162 were under mechanical ventilation at randomization. Thirty-day mortality was similar in both groups (38% in Hydroxychloroquine vs. 41% in placebo, hazard ratio [HR] 0.88, 95% Confidence Interval [95%CI] 0.51-1.53). In the surviving participants, no significant difference was found in secondary outcomes.

**Conclusion:** No beneficial effect or significant harm could be demonstrated in our randomized controlled trial including 214 patients, using relatively low doses of Hydroxychloroquine compared with placebo in hospitalized patients with severe COVID-19.

**CONSORT GUIDELINES:** 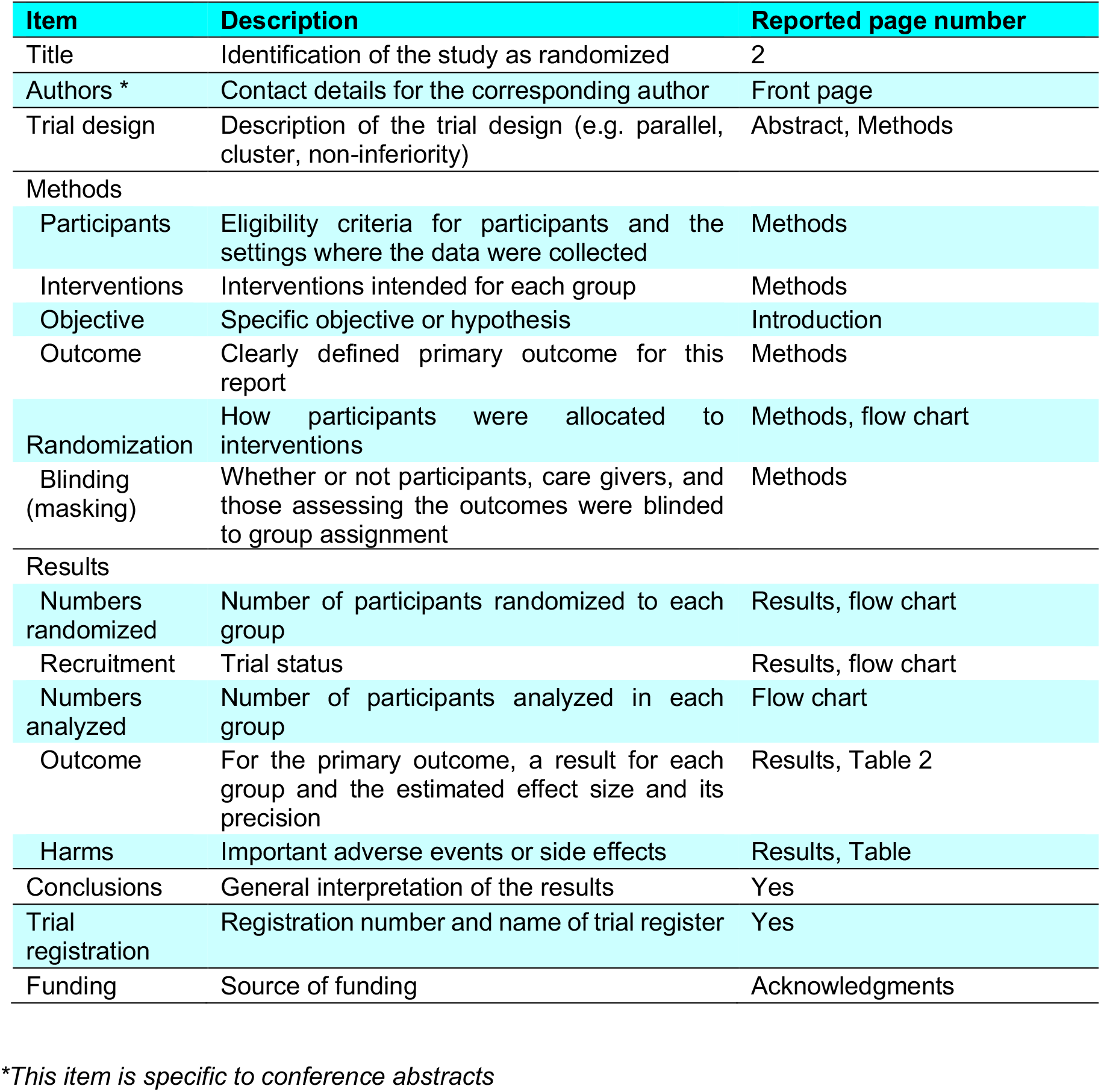

## Introduction

The outbreak of respiratory infection by the novel coronavirus 2019 (SARS-CoV-2) started in December 2019, in Wuhan (Hubei Province), China ^1-5^. From this city, the outbreak has been spreading to the majority of countries worldwide in a severe pandemic ^6^. As of July 17, 2020, more than 13 million infections and half a million deaths have been reported ^6^.

Several drugs have been prescribed for patients with COVID-19, based on their known immunomodulatory or anti-inflammatory effects, or on their *in-vitro* antiviral effects ^7^. Chloroquine and HydroxyChloroQuine (HCQ) have been in regular use for decades to treat malaria and, more recently, for the treatment of some rheumatic diseases, with a well-documented benefit/risk profile at a very low cost. The majority of published studies on HCQ or chloroquine have been observational, or relatively small controlled trials. The antiviral Remdesivir shortened time to recovery in hospitalized adults with COVID-19 compared with placebo but did not demonstrate improvement in survival ^8^. In contrast, Dexamethasone at a moderate dose had an important reduction in mortality in patients with severe COVID-19 ^9^. Several large trials on HCQ have been recently suspended, such as the RECOVERY trial ^10^, the World Health Organization SOLIDARITY trial ^11^, and an NIH-funded trial involving HCQ ^12^, but the full details of the results are unknown, as there are to our knowledge, no peer-reviewed formal publications to date.

Our aim was to estimate whether 10-day treatment with a relatively low dose of HCQ (200 mg twice daily), reduces 30-day mortality in hospitalized patients with severe COVID-19 disease, a low-cost treatment unlikely to result in important adverse effects.

## Material and methods

This was a phase III, multicenter, randomized, double-blind, parallel-control clinical trial held primarily at the Mexican National Institute of Respiratory Diseases Ismael Cosío Villegas (INER), a referral center for respiratory diseases; and at two additional participating hospitals belonging to the network of High Specialty Regional Hospitals (Hospitales Regionales de Alta Especialidad) in Ixtapaluca and Oaxaca, Mexico. All three are public hospitals that mainly treat uninsured patients and provide free treatment for COVID-19 patients. Centers differed in a) altitude above sea level, with Ixtapaluca and Mexico City sharing a similar mean altitude (2,260m and 2,240m respectively), while Oaxaca is at 1,460 m above sea level; and b) although all three centers were designated as COVID-19 hospitals, only INER was converted into a COVID-19-only referral center. This report followed the CONSORT guidelines ^13^ and was approved by the Institutional Ethics Committees. Consent to participate in the study was signed either in person or electronically by legal representatives when a participant was unable to sign due to the initiation of mechanical ventilation. The trial was registered in ClinicalTrials.gov (identifier NCT04315896).

Eligible subjects were adults over 18 years of age, with <14 days from symptoms onset, a confirmed diagnosis of COVID-19 by RT-PCR (in a pharyngeal/nasopharyngeal swab sample or in a tracheal aspirate/bronchial lavage) that required hospitalization as decided by the attending physicians. Because all patients had hypoxemia (SaO_2_ <90% ambient air or PaO_2_/FIO_2_ <250 in Mexico City) and opacities in the chest roentgenogram or the tomography, hospitalization was usually due to the requirement of respiratory support, either supplementary oxygen or mechanical ventilation. All diagnostic RT-PCR followed a similar technique (Berlin Protocol) with all laboratories standardized by the Mexican National Reference Laboratory (INDRE). RT-PCR was performed *in-situ* by participating hospitals, except in Oaxaca, where testing was performed at the state laboratory, implying a 1-2 additional delay in obtaining results and the inability to perform additional RT-PCR tests on the same patient.

We excluded patients with known previous COVID-19 infection, those previously treated with HCQ or chloroquine during the last month, pregnant woman, those with a planned transfer to another hospital unit, or those participating in another COVID-19 trial. We also excluded patients based on a contraindication to start or continue HCQ including known hypersensitivity to HCQ or chloroquine, a corrected QT interval (QTc) >0.50 s, severe liver or kidney disease, a history of pre-existing maculopathy, and to avoid to the extent possible a dangerous prolongation of the QTc and derived complications ^14^, those with >11 score points (of a maximum of 21 score points) on a scale assessing the risk of QTc prolongation in hospitalized patients, including age, gender, myocardial infarction or heart failure, sepsis, the use of drugs known to prolong the QTc or diuretics, and hypokalemia ^15^. Attending physicians were free to exclude a patient from the protocol at any time.

### Randomization

Eligible patients were randomized centrally, and separately for each participating hospital, utilizing an online-dedicated software (http://www.randomization.com), and results were employed to label flasks containing 20 tablets of the experimental drug and the identically appearing and packed sucrose placebo. Randomization considered two separate groups: (a) patients in critical condition specifically under invasive mechanical ventilation and with a disease severity grade 7-9 according to the World Health Organization (WHO) classification ^16^ with or without renal dialysis or the use of vasoactive drugs, and (b) those without invasive mechanical ventilation, with WHO classification disease severity classification of 4-6, all receiving supplementary oxygen therapy. Other categories of disease severity in the WHO classification, defined by non-invasive ventilation and the use of Extra-Corporeal Membrane Oxygenation (ECMO), were not utilized in the participating hospitals, and high-flow oxygen devices were prescribed only in 10 patients. Subjects entering the experimental group received HCQ orally or by nasogastric tube, 200 mg every 12 h, for 10 days. Subjects in the placebo group received an identical sucrose-placebo for 10 days.

### Blinding

Recruiters, patients, treating physicians, nursing staff, and the rest of the treating team, along with the follow-up evaluation monitors and the data-entry personnel, were blinded to group assignment.

### Outcomes

The main outcome was the 30-day mortality rate after randomization. Secondary outcomes included the proportion of patients requiring invasive ventilatory support after admission, duration in days of invasive mechanical ventilation for patients requiring such a procedure, duration of hospitalization in survivors, and incidence of severe adverse events leading to treatment discontinuation, intervention, or death.

Patients were treated according to the protocols of the participating institution under the responsibility of the attending physician, who could prescribe other drugs intended as a specific treatment for COVID-19, but not as part of another drug trial. Physicians-in-charge could also avoid participation in the trial if they considered the patient’s participation to be risky or inadequate or could cancel participation later in the follow-up. Monitors registered all interventions, medications for all purposes, or non-drug interventions.

Adverse events were reported regularly to the Institutional Ethics Committees and to the manufacturer of the drug and placebo (Sanofi-Aventis de México, S.A. de C.V.)

### Procedures

Each participant had a single capture form filled, with daily updates across the duration of treatment (10 days), hospital discharge, or death. If the patient was at home by day 30, the status was evaluated by a telephone call. Treatment adherence was assessed each day by counting the remaining assigned pills in the flask. In Oaxaca, a daily dose (two pills), were given daily to the nurse. All medications administered, the results of bacterial cultures, the use of supplementary oxygen or mechanical ventilation, additional support such as dialysis, vasoactive drugs, antibiotics, vital status, adverse events, the presence of a prolonged QTc (>0.5 s) from an ECG, and laboratory results were recorded each day during treatment. RT-PCR in pharyngeal or nasopharyngeal aspirate, or in tracheal aspirate/bronchial lavage if intubated, was ordered every 7 days from randomization.

### Sample Size

The original design considered, under uncertainty of a pandemic in its initial phases, a total randomized population of 600 patients (300 per group), based on an estimation of a 50% reduction in mortality from 15% in the placebo group, with a study power of 80% and a two-tailed significance alpha of 0.05. An interim analysis was planned, upon completion of one half of the sample. In mid-July, 2020, the rhythm of recruitment was reduced drastically, due to several reasons including patient refusal, that of their relatives, or that of their treating physicians, coinciding with the worldwide suspension of several large trials testing HCQ in which no benefits of the drug were found ^10-12^. Thus, it became unfeasible to complete the proposed sample size.

### Statistical analysis

Analysis was performed primarily as an intention-to-treat. We compared the 30-day death Hazard-Ratio (HR) by means of a Cox proportional-hazard model, considering right-censoring for the surviving patients. Models were fitted crude and adjusted for age, gender, SOFA score at randomization, and the number of previously diagnosed comorbidities as referred by the patient or relative (diabetes, HIV, obesity, high blood pressure, cardiovascular disease, tobacco smoking, alcoholism, asthma, Chronic Obstructive Pulmonary Disease, neurological disease, autoimmune disease, or the use of immunosuppressant’s). Similar models were fit for secondary endpoints in the survivors including a comparison of days of mechanical ventilation, days of hospitalization, time to improvement (time to extubation or time to hospital discharge), and a comparison of severe adverse events including death, but especially cardiac arrhythmia, sudden death, and QTc prolongation.

## Results

From April 8th to July 12th, 2020, 567 patients presenting at the Emergency Room (ER) were assessed; 211 did not meet inclusion criteria, 11 were included in other trials, and 131 declined to participate (see flow chart). A total of 214 patients admitted to the participating centers with clinical manifestations of COVID-19 and RT-PCR positive for SARS-COV-2, accepted to participate in the trial and were randomized as follows: 158 from INER; 18 from Ixtapaluca; and 38 from Oaxaca, 106 assigned to HCQ, and 108 to placebo (see flow chart). Two patients with a clinical and radiologic presentation consistent with COVID-19, one for each treatment group were randomized without RT-PCR confirmation and were finally reported negative for two consecutive tests. The assigned treatment was suspended, but the patients were followed, and analyzed for the main outcome. Four additional randomized patients (three in the HCQ group and one in the placebo group) did not receive the assigned treatment in the hospital ward, and one (in the placebo group) was randomized but participated in a different trial and treatment administration in our trial was suspended.

As described in **Table 1**, the mean age of the studied individuals was 49.6 ± 12 years, and the majority were males (75%). Comorbidities were present in 66%, obesity in 47%, diabetes in 16%, high blood pressure in 17%, current tobacco smoking in 11%, and cardiovascular disease in one. Median duration of symptoms before reaching the hospital was 7 days, and median days from admission to randomization were 3 days.

**TABLE 1.**
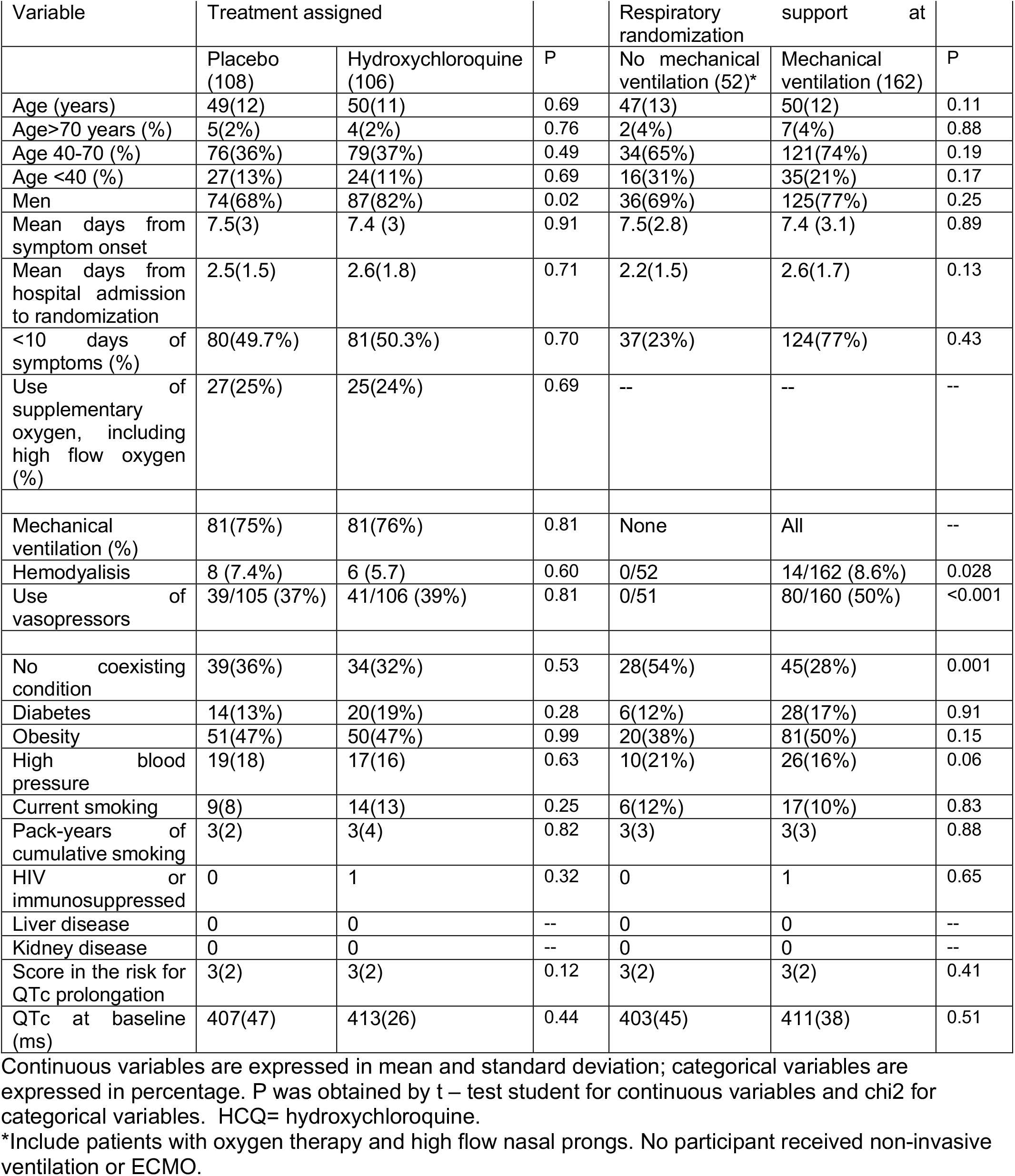
Baseline characteristics of the subjects

| Variable | Treatment assigned |  | P | Respiratory support at randomization |  | P |
| --- | --- | --- | --- | --- | --- | --- |
|  | Placebo (108) | Hydroxychloroquine (106) |  | No mechanical ventilation (52)* | Mechanical ventilation (162) |  |
| Age (years) | 49(12) | 50(11) | 0.69 | 47(13) | 50(12) | 0.11 |
| Age>70 years (%) | 5(2%) | 4(2%) | 0.76 | 2(4%) | 7(4%) | 0.88 |
| Age 40-70 (%) | 76(36%) | 79(37%) | 0.49 | 34(65%) | 121(74%) | 0.19 |
| Age <40 (%) | 27(13%) | 24(11%) | 0.69 | 16(31%) | 35(21%) | 0.17 |
| Men | 74(68%) | 87(82%) | 0.02 | 36(69%) | 125(77%) | 0.25 |
| Mean days from symptom onset | 7.5(3) | 7.4 (3) | 0.91 | 7.5(2.8) | 7.4 (3.1) | 0.89 |
| Mean days from hospital admission to randomization | 2.5(1.5) | 2.6(1.8) | 0.71 | 2.2(1.5) | 2.6(1.7) | 0.13 |
| <10 days of symptoms (%) | 80(49.7%) | 81(50.3%) | 0.70 | 37(23%) | 124(77%) | 0.43 |
| Use of supplementary oxygen, including high flow oxygen (%) | 27(25%) | 25(24%) | 0.69 | -- | -- | -- |
| Mechanical ventilation (%) | 81(75%) | 81(76%) | 0.81 | None | All | -- |
| Hemodialysis | 8 (7.4%) | 6 (5.7) | 0.60 | 0/52 | 14/162 (8.6%) | 0.028 |
| Use of vasopressors | 39/105 (37%) | 41/106 (39%) | 0.81 | 0/51 | 80/160 (50%) | <0.001 |
| No coexisting condition | 39(36%) | 34(32%) | 0.53 | 28(54%) | 45(28%) | 0.001 |
| Diabetes | 14(13%) | 20(19%) | 0.28 | 6(12%) | 28(17%) | 0.91 |
| Obesity | 51(47%) | 50(47%) | 0.99 | 20(38%) | 81(50%) | 0.15 |
| High blood pressure | 19(18) | 17(16) | 0.63 | 10(21%) | 26(16%) | 0.06 |
| Current smoking | 9(8) | 14(13) | 0.25 | 6(12%) | 17(10%) | 0.83 |
| Pack-years of cumulative smoking | 3(2) | 3(4) | 0.82 | 3(3) | 3(3) | 0.88 |
| HIV or immunosuppressed | 0 | 1 | 0.32 | 0 | 1 | 0.65 |
| Liver disease | 0 | 0 | -- | 0 | 0 | -- |
| Kidney disease | 0 | 0 | -- | 0 | 0 | -- |
| Score in the risk for QTc prolongation | 3(2) | 3(2) | 0.12 | 3(2) | 3(2) | 0.41 |
| QTc at baseline (ms) | 407(47) | 413(26) | 0.44 | 403(45) | 411(38) | 0.51 |

Continuous variables are expressed in mean and standard deviation; categorical variables are expressed in percentage. P was obtained by t – test student for continuous variables and chi2 for categorical variables. HCQ= hydroxychloroquine.
\*Include patients with oxygen therapy and high flow nasal prongs. No participant received non-invasive ventilation or ECMO.

All patients, from their first encounter in the ER, were in respiratory failure, with severe hypoxemia (mean SpO_2_ by pulse oximeter 65 ± 20%), tachycardia (pulse rate 108 ± 17 beats min^-1^), and tachypnea (breathing frequency [BF] 32 ±10 breaths min^-1^) (See Table 2). At randomization, 162 required invasive mechanical ventilation, 10 were treated with high flow oxygen, and the remaining patients received supplementary oxygen by nasal prongs (Table 2) with a mean PaO_2_/FIO_2_ of 145 ± 67 in the whole group, 130 ± 54 in those with mechanical ventilation, and 194 ± 80 in those with supplementary oxygen, but already with a slower BF (24 ± 5.9 breaths min^-1^) and heart rate (85 ± 18 beats min^-1^) compared with their arrival at the ER. Median SOFA score was 6 points in those under ventilation and 3 points in non-ventilated patients. **(Table 2)** Patients from INER more often required mechanical ventilation at randomization (86%) compared with the remaining two other participating hospitals (66% in Oaxaca, and none in Ixtapaluca, respectively).

**TABLE 2.** Key measurements at randomization.

| Variable | Treatment assigned |  |  | Respiratory support at randomization |  |  |
| --- | --- | --- | --- | --- | --- | --- |
|  | Placebo (108) | Hydroxychloroquine (106) | P | No mechanical ventilation* (52) | Mechanical ventilation (162) | P |
| **On arrival to emergency room** |  |  |  |  |  |  |
| SpO<sub>2</sub> (%) | 66 (20) | 63 (20) | 0.37 | 79(15) | 61(19) | <0.001 |
| Pulse rate (min<sup>-1</sup>) | 105 (18) | 111 (16) | 0.01 | 105(13) | 109(18) | 0.09 |
| Breathing frequency (min<sup>-1</sup>) | 32 (11) | 33 (10) | 0.58 | 27(9) | 34(10) | <0.001 |
| **On randomization** |  |  |  |  |  |  |
| PaO<sub>2</sub> (mmHg) | 69(22) | 70(11) | 0.54 | 71(31) | 69(18) | 0.60 |
| SaO<sub>2</sub> (%) | 88.2(12.7) | 90.9(9) | 0.08 | 90(10) | 90(11) | 0.70 |
| FiO<sub>2</sub> (%) | 54(22) | 56(20) | 0.45 | 39(16) | 60(20) | <0.001 |
| PaO<sub>2</sub>/FiO<sub>2</sub> (mmHg) | 148 (75) | 141 (59) | 0.42 | 207 (81) | 130 (54) | <0.001 |
| PaCO<sub>2</sub> (mmHg) | 43(15) | 44(12) | 0.78 | 35(11) | 46(13) | <0.001 |
| pH | 7.37 (0.09) | 7.37(0.10) | 0.61 | 7.43(0.06) | 7.36(0.1) | <0.001 |
| Breathing frequency (min<sup>-1</sup>) | 25 (5) | 26 (4) | 0.69 | 24(6) | 26(4) | 0.01 |
| Heart rate (min<sup>-1</sup>) | 84 (19) | 84(19) | 0.98 | 85(18) | 83(20) | 0.47 |
| Systolic Blood pressure (mmHg) | 116(16) | 116(17) | 0.94 | 116(15) | 116(17) | 0.93 |
| Diastolic Blood pressure(mmHg) | 70(12) | 70(10) | 0.81 | 72(10) | 69(11) | 0.06 |
| Creatinine (mg/dL) | 1.2(1) | 1.3(1) | 0.44 | 0.83(0.3) | 1.4(1.1) | <0.001 |
| Lymphocytes | 0.95(0.73) | 0.91(0.91) | 0.69 | 1.15(1) | 0.86(0.72) | 0.03 |
| Lactate ** | 2.2(1.5) | 2.1(1.7) | 0.85 | 1.4(0.7) | 2.4(1.7) | <0.001 |
| Glucose | 152 (69) | 167(71) | 0.14 | 132 (44) | 167(73) | 0.001 |
| C-RP** | 20(24) | 19(13) | 0.65 | 18(34) | 10(14) | 0.60 |
| Ferritin** | 1412(1483) | 1159(1210) | 0.32 | 1444(1747) | 1243(1208) | 0.48 |
| D-dimer** | 69(179) | 51(151) | 0.45 | 59(153) | 60(170) | 0.98 |
| BNP ** | 75(115) | 132(273) | 0.16 | 43(52) | 127(243) | 0.06 |
| Procalcitonin** | 8.3(49) | 7(34) | 0.84 | 0.58(1.3) | 9.7(48) | 0.26 |
| SOFA score | 5(3) | 5(2) | 0.43 | 3(2) | 6(2) | <0.001 |

Continuous variables are expressed in mean and standard deviation; categorical variables are expressed as percentage. P was obtained by the student T test for continuous variables and chi2 for categorical variables.
SpO<sub>2</sub>: oxygen saturation by pulse oxymeter; PaO<sub>2</sub>: partial pressure of oxygen in arterial blood; SaO<sub>2</sub>: arterial oxygen saturation ; FiO<sub>2</sub>: fraction of inspired oxygen ; PaO<sub>2</sub>/FiO<sub>2</sub>: ratio of arterial partial pressure of oxygen and fraction of inspired oxygen; PaCO<sub>2</sub>: partial pressure of carbon dioxide in arterial blood; C-RP: c-reactive protein; BNP: brain natriuretic peptide; SOFA: Sequential Organ Failure Assessment; HCQ: hydroxychloroquine
\*Include patients with low-flow and high-flow oxygen therapy by nasal prongs.
\*\*Values obtained in the first 3 days of the randomization

### Use of allocated treatment and suspension of treatment (see flow chart)

Seven randomized patients did not receive the allocated treatment, 4 from the HCQ group (one because a negative RT-PCR was reported twice after randomization, and three due to drug loss inside the hospital ward) and three in the placebo group (one with two negative RT-PCR reported after randomization, one due to drug loss in the hospital ward, and one who participated in another Randomized Controlled Trial (RCT). The randomized treatment was received for a mean of 8 days (SD 3) without difference between the two groups (mean number of pills taken from the bottle 16±6). Randomized groups were well-matched (Tables 1 and 2), but most recruited patients (76%) required mechanical ventilation at the time of randomization. The HCQ group had more males (82% vs. 68%, p = 0.02) but a similar SOFA score, PaO_2_/FIO_2_, blood pressure and creatinine upon hospital admission (Table 2). All surviving patients completed the 30-day follow-up.

### Use of other medications

Use of other medications during the 10-day treatment of HCQ or placebo, was very common: Clarithromycin was prescribed to 146 (68%) patients, and Azithromycin to 50 (23%) of patients, prescribed usually as part of the antibiotic coverage of suspected bacterial pneumonia. A cephalosporin was prescribed to 182 (85%), a carbapenem to 182 (85%), Oseltamivir to 39 (18%), Lopinavir/Ritonavir to 62 (29%), and anticoagulants to 120 (56%). Tocilizumab was not available at the hospitals and was prescribed only in five cases.

Systemic corticosteroids were prescribed to 114 subjects, more than 50% of the study population; the most frequent systemic corticosteroid was Methylprednisolone followed by Dexamethasone without a difference among the treatment groups. Patients who required mechanical ventilation were more frequently prescribed a systemic corticosteroid. **(Table 3**.) Methylprednisolone doses varied, but on average were 100 ± 72 mg per day; Dexamethasone doses were 5 ± 2 mg per day.

**TABLE 3.**
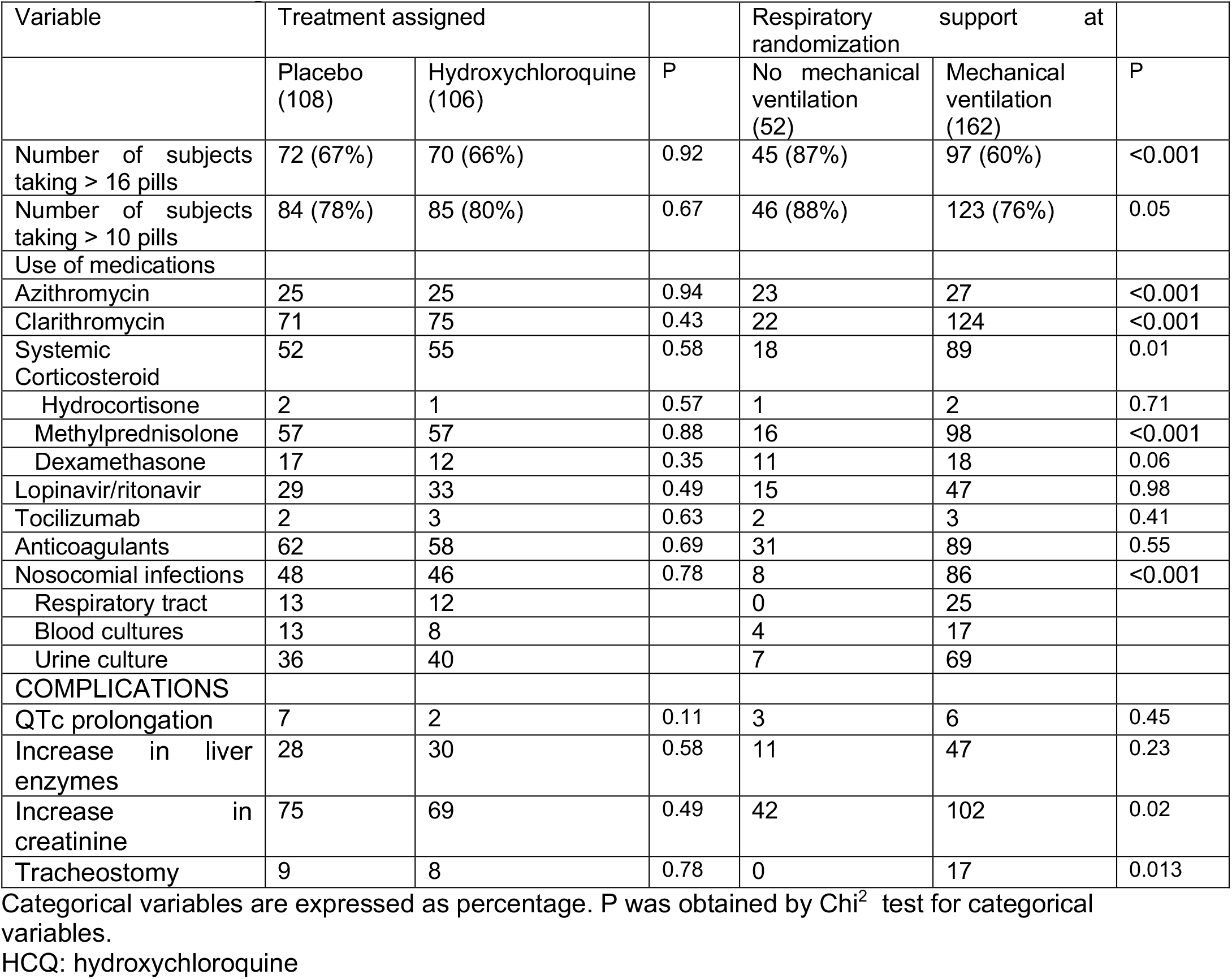
Coexisting medications and adverse events

| Variable | Treatment assigned |  |  | Respiratory support at randomization |  |  |
| --- | --- | --- | --- | --- | --- | --- |
|  | Placebo (108) | Hydroxychloroquine (106) | P | No mechanical ventilation (52) | Mechanical ventilation (162) | P |
| Number of subjects taking > 16 pills | 72 (67%) | 70 (66%) | 0.92 | 45 (87%) | 97 (60%) | <0.001 |
| Number of subjects taking > 10 pills | 84 (78%) | 85 (80%) | 0.67 | 46 (88%) | 123 (76%) | 0.05 |
| Use of medications |  |  |  |  |  |  |
| Azithromycin | 25 | 25 | 0.94 | 23 | 27 | <0.001 |
| Clarithromycin | 71 | 75 | 0.43 | 22 | 124 | <0.001 |
| Systemic Corticosteroid | 52 | 55 | 0.58 | 18 | 89 | 0.01 |
| Hydrocortisone | 2 | 1 | 0.57 | 1 | 2 | 0.71 |
| Methylprednisolone | 57 | 57 | 0.88 | 16 | 98 | <0.001 |
| Dexamethasone | 17 | 12 | 0.35 | 11 | 18 | 0.06 |
| Lopinavir/ritonavir | 29 | 33 | 0.49 | 15 | 47 | 0.98 |
| Tocilizumab | 2 | 3 | 0.63 | 2 | 3 | 0.41 |
| Anticoagulants | 62 | 58 | 0.69 | 31 | 89 | 0.55 |
| Nosocomial infections | 48 | 46 | 0.78 | 8 | 86 | <0.001 |
| Respiratory tract | 13 | 12 |  | 0 | 25 |  |
| Blood cultures | 13 | 8 |  | 4 | 17 |  |
| Urine culture | 36 | 40 |  | 7 | 69 |  |
| COMPLICATIONS |  |  |  |  |  |  |
| QTc prolongation | 7 | 2 | 0.11 | 3 | 6 | 0.45 |
| Increase in liver enzymes | 28 | 30 | 0.58 | 11 | 47 | 0.23 |
| Increase in creatinine | 75 | 69 | 0.49 | 42 | 102 | 0.02 |
| Tracheostomy | 9 | 8 | 0.78 | 0 | 17 | 0.013 |

Categorical variables are expressed as percentage. P was obtained by Chi<sup>2</sup> test for categorical variables.
HCQ: hydroxychloroquine

### Outcomes

Among all participating individuals, 39% died: 47% of those requiring mechanical ventilation, and 13% of the remaining participants. No significant difference in the main outcome, 30-day mortality, could be identified in treatment groups (38% in HCQ, 41% in placebo, HR 0.89, and 95%CI 0.58-1.38), either in all participants, **(Figure 1 and 2)** or on separating those with mechanical ventilation and non-ventilated patients with supplementary oxygen. **Figure 2** depicts the impact of treatment on mortality, according to the level of respiratory support (mechanical ventilation or supplementary oxygen) and recruitment site. Results are considerably influenced by the most numerous number of patients being from Mexico City, with most requiring mechanical ventilation at randomization. The Relative Risk (RR) of death with HCQ was 0.43 (95%CI 0.09-2.03) in non-ventilated patients, comprising about one quarter of the recruited patients, and was 0.20 in Ixtapaluca (95%CI 0.03-1.39), a site recruiting only 18 patients.

**FIGURE 1.**
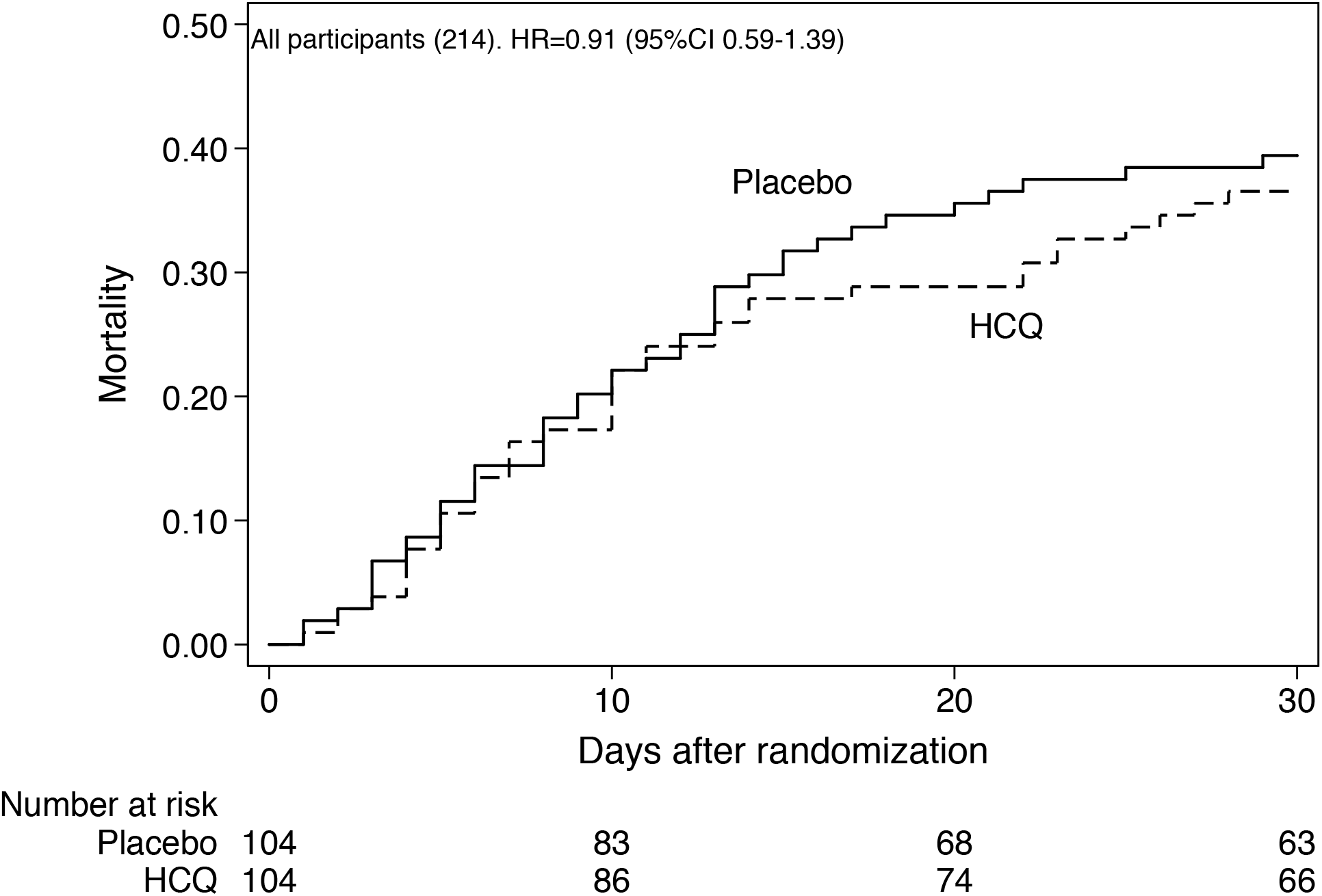
Death in all participants, hydroxychloroquine (HCQ) group vs. placebo. Unadjusted mortality graph, Placebo (continuous line) vs HCQ (dashed line).

**FIGURE 2.**
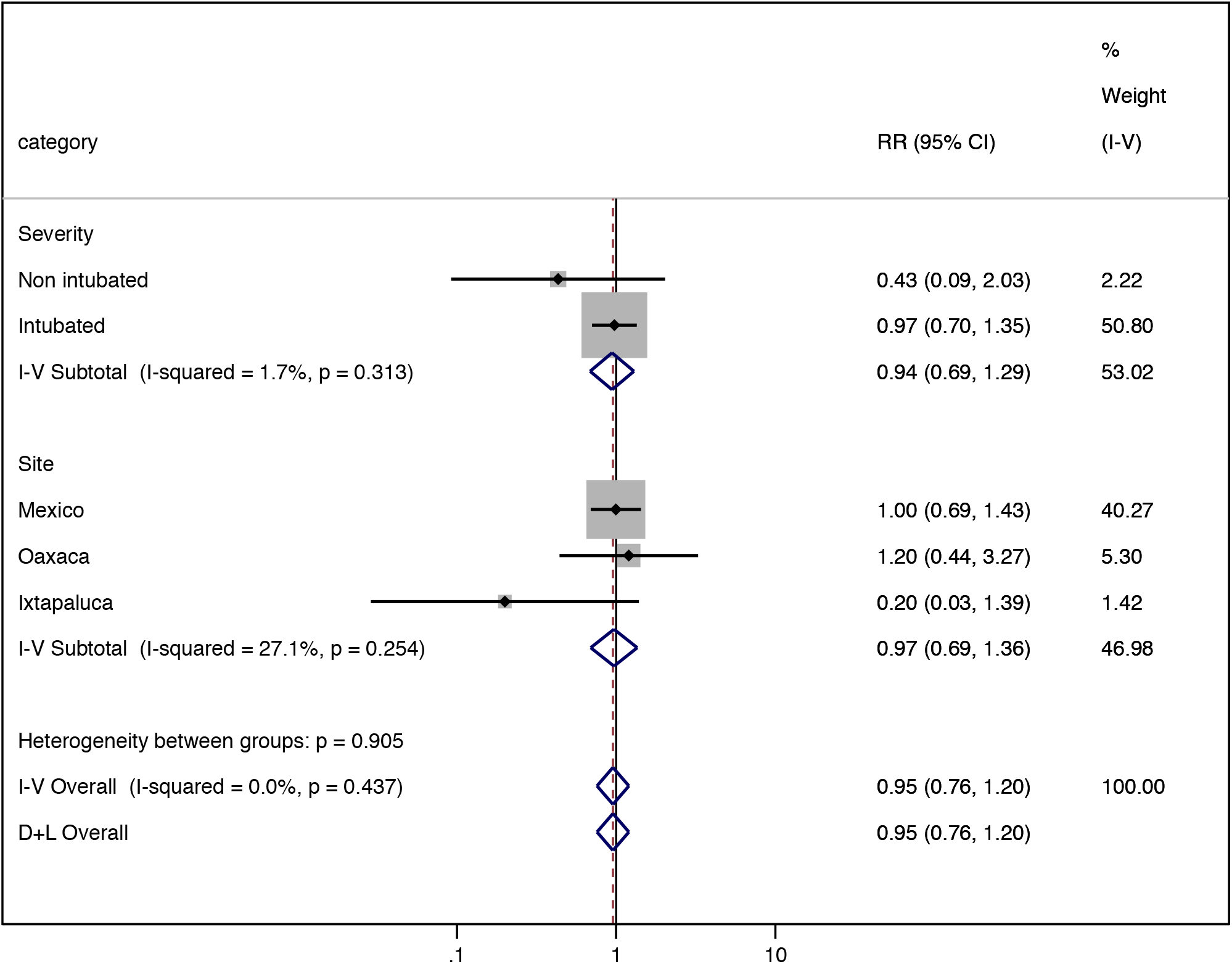
Mortality by mechanical ventilation and site. Impact of HCQ in relevant subgroups, by severity of respiratory failure (intubated vs non intubated) and by recruitment site. Heterogeneity was I^2^ p=0.905. 0.95 (0.76-1.20).

Secondary outcomes were not significantly different between treatment groups: in terms of days of mechanical ventilation (13.8 ± 12 days in HCQ vs. 14.0 ± 12 days in placebo, HR 0.98, 95% CI 0.64-1.52), days of hospitalization (17.8 ± 11 days in HCQ vs. 18.1 ± 12 days in the placebo group, HR 1.01, 95% CI 0.72-1.43), or time to improvement (14.2 ± 10 days in HCQ vs. 15.2 ± 10 days in the placebo group, HR 1.05, 95% CI 0.75-1.49). **(Table 4)**

**TABLE 4.** OUTCOMES OF THE TRIAL n/total patients (%) or means (SD)

| Outcome | Treatment groups |  | Crude Hazard Ratio<br>(95%CI) | Adjusted Hazard Ratio<br>(95%CI)* |
| --- | --- | --- | --- | --- |
|  | Placebo<br>(108) | Hydroxychloroquine<br>(106) |  |  |
| Primary Outcome |  |  |  |  |
| Mortality at 30 days<sup>1</sup> | 44/108 (41%) | 40/106 (38%) | HR 0.91 (0.59-1.39) | HR 0.80 (0.51-1.23) |
| Secondary outcomes |  |  |  |  |
| Days at hospital in survivors | 18.1 (12) | 17.8 (11) | 1.00 (0.71-1.42) | 1.37 (0.95-1.97) |
| Days on mechanical ventilation in survivors and with mechanical ventilation | 14.0(12) | 13.8(12) | 0.93 (0.57-1.52) | 1.07 (0.62-1.83) |
| Improvement (days to extubation in ventilated or exit from hospital in survivors<sup>2,3</sup> | 15.2 (10) | 14.2 (10) | 1.07 (0.76-1.51) | 1.35 (0.94-1.95) |
| Death or other severe adverse events <sup>4</sup> | 54/108 (50%) | 55/106 (52%) | 0.95 (0.65-1.40) | 0.87 (0.59-1.28) |

HCQ= hydroxychloroquine, \* Models are adjusted by age, gender, comorbidities, SOFA, site and requirement of mechanical ventilation at randomization regression model. Effect size (Cohen's d) in days at hospital (0.03), days on mechanical ventilation (0.02) and days to improvement (0.1), proportion with severe adverse events (0.04), or deaths (0.06) were very small.

### Adverse Events

No significant difference in severe adverse events, including deaths was observed in the treatment groups (52% in HCQ, vs. 54% in placebo, HR 0.95, 95% CI 0.65-1.40) (**Table 3**). Eight patients had treatment discontinued due to an adverse event (four in each group), and in 20, the attending physician discontinued treatment (eight in the HCQ group and 12 in the placebo group). Qtc prolongation (>0.5 s) was identified in nine subjects and in six, it was reported as a severe adverse event. Nevertheless, no significant difference was found between the treatment groups. Increase in serum creatinine levels was one of the most frequently reported adverse events, without difference between treatment groups: 144 subjects had an increase in serum creatinine of >1.3 times the upper limit of normal of the clinical laboratory, and in 40 subjects it was reported as a severe adverse event, with 14 of these patients treated with hemodialysis.

Significant predictors of mortality were the following: (a) requirement of intubation and mechanical ventilation (HR 4.3, 95%CI 2.0-9.2); (b) masculine gender (HR 1.7, 95% CI 9.96-2.92); (c) age>65 years (HR 1.67, 95% CI 0.91-3.1); (d) number of reported comorbidities (HR 1.65, 95% CI 1.2-2.2); (e) obesity (HR 1.64, 95% CI 1.1-2.5); (f) body mass index (HR 1.06, 95% CI 1.03-1.10); and (g) SOFA score (HR 1.2, 95% CI 1.1-1.3). On including all previous variables in a multivariate Cox proportional hazard model, the requirement of invasive mechanical ventilation (HR 3.9, 95%CI 1.8-8.4), male sex (HR 1.8, 95% CI 1.02-3.1), body mass index (HR 1.07, 95% CI 1.02-1.12), and number of comorbidities (HR 1.49, 95%CI 1.02-2.2), remained significant predictors of in-hospital death. In a Cox model including age, sex, number of comorbidities, SOFA score, mechanical ventilation, and site, two variables considerably increased the risk of death: mechanical ventilation at randomization (HR 16.4, 95%CI 2.2-121), and recruitment in Ixtapaluca (HR 12.6 95%CI 1.5-106). Patients from Ixtapaluca comprised a small group (n=18), none requiring mechanical ventilation at randomization but 4/6 patients who subsequently died required mechanical ventilation within a short time after randomization.

### Nosocomial infections

During the 30-day follow-up, 94 patients (46%) were reported with a health-related infection, 47 in the placebo group and 45 in the HCQ group, 86/92 patients were on mechanical ventilation, and the remaining patients were on supplementary oxygen. Sources of bacterial growth in culture were from airways secretions (20%), from blood (16%), and from urine (54%). The isolated bacteria were 13% *Staphylococcus spp*., 26% *Pseudomonas aeruginosa*, 4% *Stenotrophomonas maltophilia*, 14% *Acinetobacter baumanni*, 19% *Klebsiella pneumoniae, Streptococcus pneumoniae* 1%, 9% *Candida spp*., 5% *Enterobacter spp*., and, 16% *Escherichia coli*. No difference by treatment group was found in the bacteria isolated except in the *Acinetobacter* present in 10 individuals of the placebo group and in three of the HCQ group (*p* = 0.045). **(Table 5)** The presence of nosocomial infection (any type or respiratory), co-treatment with Azithromycin or other antibiotics, administration of systemic steroids, hemodialysis, or the requirement of vasopressors did not increase the risk of death once mechanical ventilation, age, gender, SOFA score, and comorbidities were taken into account.

**TABLE 5.** Isolated microorganisms in different clinical samples.

| Bacteria | Respiratory tract | Blood | Urine |
| --- | --- | --- | --- |
| *Pseudomonas Aeruginosa* | 8(42%) | 1(7%) | 13(25%) |
| *Klebsiella pneumonie* | 8(42%) | 1(7%) | 6(12%) |
| *Escherichia coli* | 4(21%) | 1(7%) | 9(18%) |
| *Acinetobacter baumannii* | 2(10%) | 1(7%) | 9(18%) |
| *Staphylococcus Aureus* | 1(5%) | 3(20%) | 7(14%) |
| *Candida sp* | 1(5%) | 0 | 7(14%) |
| *Streptococcus pneumoniae* | 1(5%) | 0 | 0 |
| *Enterobacter* | 0 | 1(7%) | 4(8%) |

\*Except for *Acinetobacter* (more common in placebo), no differences were found between placebo and HCQ (10 in placebo and 3 in HCQ group, $p=0.045$ )

## Discussion

In a randomized trial compared with placebo, HCQ did not significantly reduce the 30-day mortality of an especially severe and hypoxemic group of patients with COVID-19. Secondary outcomes also failed to be improved by HCQ, but importantly severe toxicity was not observed to a greater degree in treated patients, despite the concomitant use of other drugs, including Azithromycin. Azithromycin was commonly prescribed as part of an empiric antibiotic for pneumonia, but also, especially at the beginning of the pandemic in Mexico, as an attempt to modify the natural course of the COVID-19.

Risk factors for death in our group included age, masculine gender, SOFA score, body mass index, and the need for mechanical ventilation, the latter found to be the most significant. Mortality of the studied population was very high compared with severe patients at our own hospitals prior to COVID-19, but the majority of our patients were in respiratory failure with mechanical ventilation, arriving with severe hypoxemia (mean SpO_2_ 63%), requiring vasopressor support, and after several days of symptoms (7 days on average).

All three hospitals participating in the trial possessed important surge-capacity preparations for COVID-19. Since the epidemic developed in China, all three hospitals aimed at preparing a larger number of beds with access to mechanical ventilation, finally reaching around three times the original number of beds available. The National Institute of Respiratory Diseases has a total of 178 beds in seven wards devoted to respiratory diseases, and 30 beds with a ventilator in the Intensive Care Unit, and in the ER. The Institute was transformed into a hospital with >100 beds with access to mechanical ventilation distributed in all wards. However, trained personnel for that number of intensive care beds was scarce, and scarcity increased because of COVID-19 infections among the personnel, and especially after a presidential decree that sent home all workers more than 65 years of age or with comorbidities.

Even though new personnel was hired, the majority were recently graduated physicians, nurses, and allied health personnel with little experience with critical patients. With the preparation of the hospital, the capacity of the mechanical ventilation services was not overwhelmed as occurred in other countries before. Instead, limitations derived from insufficient personnel with proper training in intensive care, and the occasional scarcity of medicines, and Personal Protection Equipment. The frequency of nosocomial infection was high, although in Cox proportional hazard models the presence of these nosocomial infections did not increase mortality once mechanical ventilation was taken into account.

Although this was a trial with proper randomization and blinding, demonstrated by the comparison of baseline characteristics of the treatment arms, reducing the possibility of biases due to known or unknown variables, the trial ended short of the planned sample size. Increasing the number of recruited patients proved very difficult with a growing number of refusals by patients, relatives, and treating physicians once the large trials, including RECOVERY, SOLIDARITY, and that supported by NIH, suspended their treatment arms with HCQ due to a lack of beneficial effect, although no harm from HCQ was reported. Information of these suspended trials traveled by newspapers and media ^17^, and reached the widespread population with a great impact, even before a proper peer-reviewed publication was available and analyzed, because of the considerable prestige and importance of the institutions responsible for the trials. It is understandable that in the middle of a pandemic, a rapid presentation of results of large, proper clinical trials may help to select the best treatments to improve patients or avoid drugs lacking benefit or generating harm but, on the other hand, it lead to the premature termination of several trials.

As a consequence, the sample size recruited is small and unfortunately, it is possible to miss relevant beneficial or harmful effects. Even though the best estimate of the impact of HCQ on mortality was 0.88, the confidence interval was from 0.51, implying a reduction of mortality to one half of that observed in the placebo group, to 1.53, a 50% increase in mortality compared with the placebo group. Sample size (214) had only a 80% study power to detect a relative risk of dying of 0.57 (from 41% in the control group to 23% in the HCQ group) or an RR of 1.46 (from 41% in the control group to 60% in the HCQ group) in the case of HCQ being harmful. Notwithstanding this, it can be argued that once the information of large trials showed no benefit for hospitalized patients but also no clear harm, both possibilities are unlikely. For a possible participant interested in our trial and other similar trials, once a relevant benefit in large Randomized Controlled Trials (RCT) was ruled out with reasonable certainty, the balance shifted towards risks and side effects and it became questionable to continue the trial. Dozens of trials including HCQ as treatment were registered in Clinical Trials, and it is likely that several are going to end up short, as ours did. Nonetheless, and fortunately, information can be compiled later in meta-analyses and systematic analyses.

We and others could demonstrate that HCQ side effects can be minimized with proper follow-up keeping track of the QTc segment and utilizing instruments such as the multivariable Tisdale’s scale score ^15^ to predict individuals at higher risk of QTc prolongation and its complications, combined with a relatively low dose of HCQ, safe even for prolonged periods for the majority of patients, and lacking a loading dose. Our population was using different types of medications including Azithromycin, several antibiotics, systemic corticosteroids, and Lopinavir/Ritonavir, in an attempt to improve survival, the majority of the time before any drug demonstrated improvement of patients with COVID-19.

## Conclusions

In summary, no beneficial effect or significant harm could be demonstrated in our randomized controlled trial including 214 patients, using relatively low doses of HCQ compared with placebo in hospitalized patients with severe COVID-19. However, the study was stopped early and likely was underpowered for finding a statistically and clinically important difference in the primary outcome.

## Data Availability

Data and Code Availability
The data and codes related to the findings of this study will be available from the corresponding author after publication, upon reasonable request, especially from investigators compiling hydroxychloroquine studies to treat COVID-19. The group formed by the principal investigators will analyze the quality of the proposal and the security offered for the data. The patient-level data, without individual identifiers, will be shared after approval of the submitted proposal.

## ACKNOWLEDGMENTS

We thank all the study participants. Support for the trial was received from the participating hospitals: Instituto Nacional de Enfermedades Respiratorias, Hospital Regional de Alta Especialidad de Ixtapaluca and Hospital de Regional de Alta Especialidad de Oaxaca, and from CONACYT (National Council of Science and Technology of Mexico) and from SANOFI through an investigator-sponsored trial, including the tested drug and identical placebo. The authors designed the trial, performed the analysis and wrote the manuscript.

---

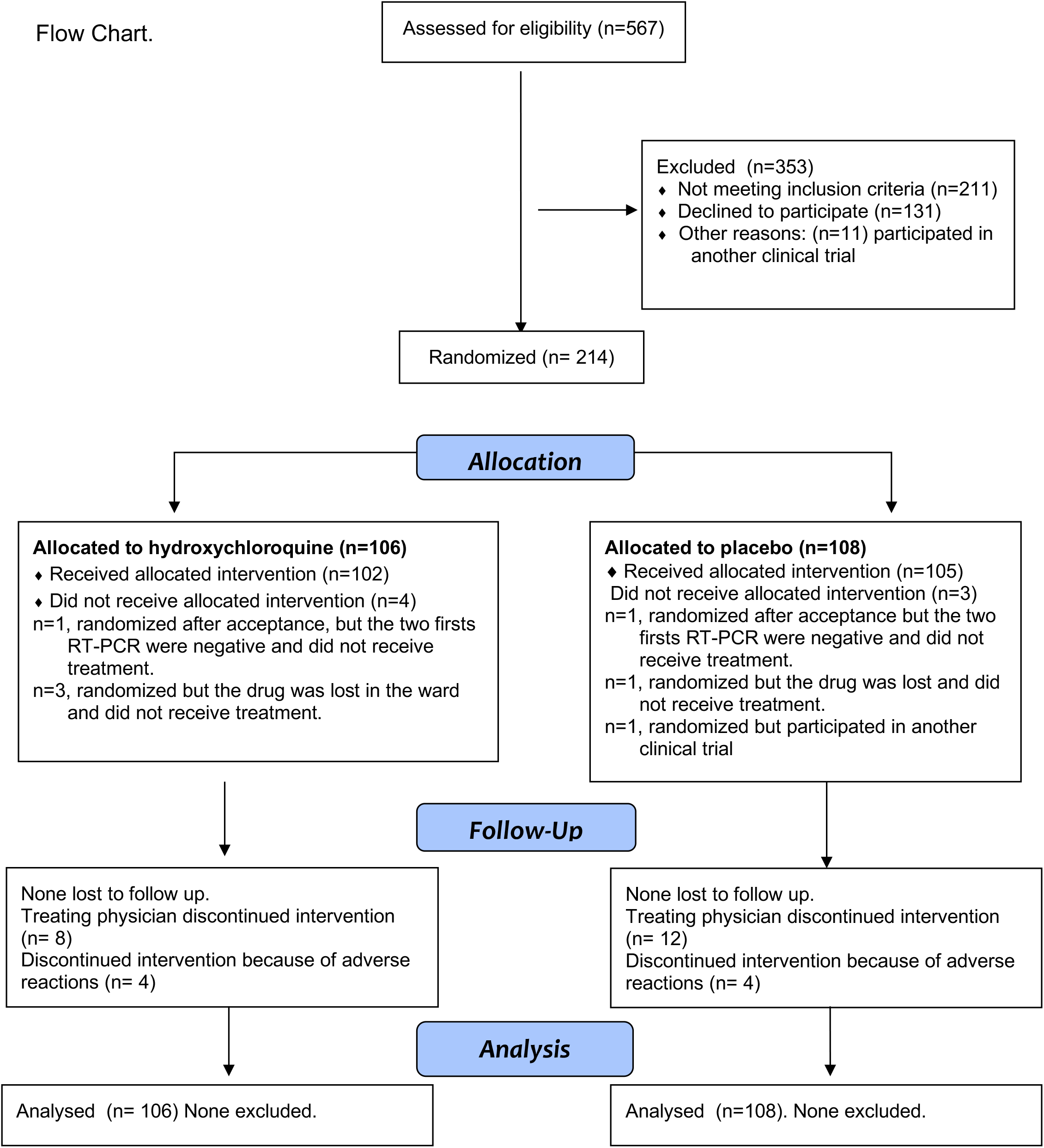

## REFERENCES

1. Chan JF, Yuan S, Kok KH, et al. A familial cluster of pneumonia associated with the 2019 novel coronavirus indicating person-to-person transmission: a study of a family cluster. Lancet. 2020;395(10223):514–523.

2. Chen N, Zhou M, Dong X, et al. Epidemiological and clinical characteristics of 99 cases of 2019 novel coronavirus pneumonia in Wuhan, China: a descriptive study. Lancet. 2020.

3. Holshue ML, DeBolt C, Lindquist S, et al. First Case of 2019 Novel Coronavirus in the United States. The New England journal of medicine. 2020;382(10):929–936.

4. Zhou P, Yang XL, Wang XG, et al. A pneumonia outbreak associated with a new coronavirus of probable bat origin. Nature. 2020;579(7798):270–273.

5. Zhu N, Zhang D, Wang W, et al. A Novel Coronavirus from Patients with Pneumonia in China, 2019. The New England journal of medicine. 2020;382(8):727–733.

6. World Health Organization. Coronavirus disease (COVID-19) Situation Report–179. 2020; https://www.who.int/docs/default-source/coronaviruse/situation-reports/20200717-covid-19-sitrep-179.pdf?sfvrsn=2f1599fa_2. Accessed July 18, 2020.

7. Wang M, Cao R, Zhang L, et al. Remdesivir and chloroquine effectively inhibit the recently emerged novel coronavirus (2019-nCoV) in vitro. Cell research. 2020;30(3):269–271.

8. Beigel JH, Tomashek KM, Dodd LE, et al. Remdesivir for the Treatment of Covid-19 - Preliminary Report. The New England journal of medicine. 2020(May 22. doi: 10.1056/NEJMoa2007764.).

9. Recovery Collaborative Group, Horby P, Lim WS, et al. Dexamethasone in Hospitalized Patients with Covid-19 - Preliminary Report. The New England journal of medicine. 2020.

10. Randomised Evaluation of COVid-19 thERapY (RECOVERY) Trial. No clinical benefit from use of hydroxychloroquine in hospitalised patients with COVID-19. 2020; https://www.recoverytrial.net/news/statement-from-the-chief-investigators-of-the-randomised-evaluation-of-covid-19-therapy-recovery-trial-on-hydroxychloroquine-5-june-2020-no-clinical-benefit-from-use-of-hydroxychloroquine-in-hospitalised-patients-with-covid-19.

11. World Health Organization. WHO discontinues hydroxychloroquine and lopinavir/ritonavir treatment arms for COVID-19. 2020; https://www.who.int/news-room/detail/04-07-2020-who-discontinues-hydroxychloroquine-and-lopinavir-ritonavir-treatment-arms-for-covid-19.

12. National Institutes of Health. NIH halts clinical trial of hydroxychloroquine. 2020; https://www.nih.gov/news-events/news-releases/nih-halts-clinical-trial-hydroxychloroquine. Accessed June 20, 2020.

13. Moher D, Hopewell S, Schulz KF, et al. CONSORT 2010 Explanation and Elaboration: updated guidelines for reporting parallel group randomised trials. The BMJ. 2010;340.

14. Mercuro NJ, Yen CF, Shim DJ, et al. Risk of QT Interval Prolongation Associated With Use of Hydroxychloroquine With or Without Concomitant Azithromycin Among Hospitalized Patients Testing Positive for Coronavirus Disease 2019 (COVID-19). JAMA cardiology. 2020;5(9):1036.

15. Tisdale JE, Jaynes HA, Kingery JR, et al. Development and validation of a risk score to predict QT interval prolongation in hospitalized patients. Circulation. Cardiovascular quality and outcomes. 2013;6(4):479–487.

16. Marshall JC, Murthy S, Diaz J, et al. A minimal common outcome measure set for COVID-19 clinical research. The Lancet Infectious Diseases. 2020;20(8):e192–e197.

17. Saitz R, Schwitzer G. Communicating Science in the Time of a Pandemic. JAMA,. 2020;324(5):443.

